# Development and validation of blood-based predictive biomarkers for response to PD-(L)-1 checkpoint inhibitors: evidence of a universal systemic core of 3D immunogenetic profiling across multiple oncological indications

**DOI:** 10.1101/2021.12.21.21268094

**Authors:** Ewan Hunter, Mehrnoush Dezfouli, Christina Koutsothanasi, Adam Wilson, Francisco C. Santos, Matthew Salter, Jurjen W. Westra, Ryan Powell, Ann Dring, Benedict Egan, Matthew Parnall, Morgan Thacker, Jayne Green, Aroul Ramadass, Serene Ng, Chun Ren Lim, Cheah Soon Keat, Ang Tick Suan, Rakesh Raman, Ho Kean Fatt, Fabian Lee Wei Luen, Thomas Guiel, Robert Heaton, Jedd Levine, Alexandre Akoulitchev

**Affiliations:** Oxford BioDynamics Plc, Oxford, UK; Oxford BioDynamics (M) Sdn Bhd, Penang, Malaysia; Mount Miriam Cancer Hospital (MMCH), Penang, Malaysia; Island Hospital, Penang, Malaysia; Oxford BioDynamics Inc., Gaithersburg, MD, USA

## Abstract

Unprecedented advantages in cancer treatment with immune checkpoint inhibitors (ICI) remain limited to a subset of patients. Systemic analyses of the regulatory 3D genome architecture linked to individual epigenetics and immunogenetic controls associated with tumour immune evasion mechanisms and immune checkpoint pathways reveals a highly prevalent patient molecular profiles predictive of response to PD-(L)1 immune checkpoint inhibitors. A clinical blood test based on the set of 8 3D genomic biomarkers has been developed and validated on several independent cancer patient cohorts to predict response to PD-(L)1 immune checkpoint inhibition. The predictive 8 biomarker set is derived from prospective observational clinical trials, representing 229 treatments with Pembrolizumab, Atezolizumab, Durvalumab, in diverse indications: melanoma, non-small cell lung, urethral, hepatocellular, bladder, prostate cancer, head and neck, vulvar, colon, breast, bone, brain, lymphoma, larynx cancer, and cervix cancers.

The 3D genomic 8 biomarker panel for response to immune checkpoint therapy achieved high accuracy up to 85%, sensitivity of 93% and specificity of 82%. This study demonstrates that a 3D genomic approach could be used to develop a predictive clinical assay for response to PD-(L)1 checkpoint inhibition in cancer patients.

## Background

Insights into tumour immunology related to mechanisms of tumour immunosurveillance and anti-tumour immune responses have led to unprecedented advances in cancer treatment. This breakthrough in cancer treatment takes advantage of key mechanisms that mitigate tumour immune evasion by targeting immune checkpoints to enhance anti-tumour immunity and harnesses overall patient immune system. These agents have shown considerable clinical benefit in certain patient populations [1–3].

Under physiological conditions immune checkpoint molecules regulate the immune system by dampening the immune response following successful mitigation of an infection and preventing the onset auto-immune conditions. One of the first identified inhibitory checkpoints - CD152, also known as T lymphocyte-associated protein (CTLA-4), has been shown to prevent expansion of CD4+ helper T cells, boost regulatory T cells, and promote a pro-tumour immuno-suppressive phenotype [4,5]. Strategies to antagonize CTLA-4 as means of increasing anti-tumour immunity, eventually lead to US Food and Drug Administration (FDA) approval of ipilimumab for treatment of metastatic melanoma [6]. With limited other therapeutic options to improve survival of advanced melanoma patients, ipilimumab demonstrated 2-year survival rate of 23.5%. However, consistent with auto-immunity observed in pre-clinical models targeting CTLA-4 [7,8], treatment with ipilimumab was associated with immune-related adverse effects in 60% of patients [9].

Today the two most successfully exploited immune checkpoints are CD279, called programmed cell death protein 1 (PD-1), expressed on tumour infiltrating lymphocytes, B cells, NK cells and myeloid cells; and its ligand CD274, called programmed death-ligand 1 (PD-L1), expressed on tumour cells. The PD-L1/PD-1 interaction is a major mechanism leading to tumour immune evasion. Agents that interfere with this interaction have demonstrated potent and durable anti-tumour activities, with less severe immune related toxicity compared to CTLA-4 blockade [3]. Accordingly, in earlier clinical trials the anti-PD-1 antibodies nivolumab and pembrolizumab have been already proven effective in treatments of melanoma, non-small cell lung cancer (NSCLC), and colorectal cancer [10–14], while anti-PD-L1 antibodies, such as atezolizumab, avelumab, durvalumab – in treatments of NSCLC, urothelial carcinoma, triple negative breast cancer [15–18]. Additional cancer types are currently under active clinical investigation.

In addition to the expanding array of therapeutic agents targeting PD-(L)1 (dostarlimab, tyvyt, libtayo, tislelizumab, camrelizumab and sasanlimab), a series of novel immune checkpoint molecules are undergoing evaluation (LAG-3/CD223, TIM-3, TIGIT, VISTA, B7-H3/CD276, BTLA/CD272) [3].

Limitations of currently approved immune checkpoint inhibitors (ICIs) include variable responses among cancer types, primary resistance in the majority of patients with objective responses observed in the minority, acquired resistance in most cancer types, and significant risk of immune related adverse side effects. The objective response rate (ORR) for anti-CTLA-4 ipilimumab (Yervoy) in melanoma was 10.9%, with high-grade treatment-related adverse event rate of 15% [9]; for anti-PD-1 pembrolizumab (Keytruda) in advanced melanoma – 33% ORR, with 14% high-grade treatment related adverse events [19]; for anti-PD-L1 avelumab (Bavenco) in urothelial carcinoma 17% ORR, with 8% high-grade treatment adverse events [20].

In practical terms, oncologists prescribing an ICI must weigh the risk of immune-related adverse effects against the ORR and benefits of ICI treatment, with very limited data to guide the decision. This has led to multiple efforts to develop predictive biomarkers to identify patients who will benefit from treatment. The predictive value of tumour intrinsic factors such as tumour mutational burden (TMB), microsatellite instability (MSI) and DNA mismatch repair deficiencies (dMMR) has been supported by several studies [21–23]. In 2017 the FDA approved pembrolizumab for treatment of advanced paediatric and adult solid tumours with high MSI and dMMR, which have not responded to prior treatments and have no other alternative treatment options [3]. The association of genetic biomarkers such as TMB with response to ICI treatment is not observed in all patients [21], and has been reported to have limited predictive value in particular study context [24].

Also, the advanced technology and standardization required for such biopsy-based tests impose practical limitations on applicability of these biomarkers in practice-based clinical settings.

Assessment of tumour infiltrating lymphocytes has been evaluated for predictive value with mixed success [25], but standardization of histologic evaluation may improve reliability [26].

Earlier trials of anti-PD-(L)1 inhibitors reported association between tumour PD-L1 expression and response to treatment in melanoma and NSCLC [27,28]. In contrast, other reports showed that durable responses to ICI could be obtained in the absence of tumour PD-L1 expression [29]. The variability in the definitions of PD-L1 positivity and in methods of its evaluation account for significant inconsistencies in predictive results, with calls for further standardization [3]. The 3D configuration of the genome plays a crucial role in coordinated gene regulation and homeostasis of cellular phenotype [30–32]. Three-dimensional genome architecture has been shown to act as the regulatory interface and integration point for genetic risks, epigenetic cues and modifications, metabolic signalling, and transcriptional events all integrated into manifestation of specific cellular phenotypes and, ultimately, clinical outcomes [30,33,34]. *EpiSwitch®* is a biomarker platform and methodology for patient stratification developed on the basis of the original chromosome conformation capture (3C) approach as a novel biomarker modality [35,36]. *EpiSwitch®* platform has reduced to practice all stages of the discovery, development, validation and monitoring of blood-based biomarkers, based on 3D genome architecture. To date, 3D genomic *EpiSwitch®* biomarkers, also known as chromosome conformation signatures (CCSs); have been used in blood test format to stratify melanoma patients; prognostically stratify patients with fast versus slow progressing motor neurone disease; stratify patients with symptomatic and pre-symptomatic neurodegenerative disease; diagnose patients with thyroid cancer and various stages of prostate cancer; prognostically stratify patients for outcome in diffuse large B cell lymphoma; predictively stratify patients with non-small cell lung cancer for response to the anti-PD-L1 immune checkpoint inhibitor avelumab; prognostically stratify high-risk individuals with immune-health profile susceptible to systemic hyperreaction and severe COVID disease complications upon infection with SARS-CoV-2 [37–52].

Based on predictive and prognostic methodologies developed with systemic 3D genomic biomarkers, we looked at the *EpiSwitch®* platform 3D genomic profiles in patients treated with immune checkpoint inhibitors to see if any could be used to predict responsiveness to ICI treatment. We have focused on the PD-(L)1 pathway target, as it is the most advanced in terms of clinically developed inhibitors. We have based our analysis on a prospective observational study with the use of several of the approved ICIs for a variety of oncological indications.

We have used the EpiSwitch® Explorer array platform for whole genome profiling of patients prior to ICI treatment, with subsequent classification of clinical outcome of response based on standard ORR criteria/ RECIST 1.1, as a standard in clinical practice and trials settings [53,54]. After analysing 1.1 million data points with annotations across the whole genome for each screened patient, we identified significant and reproducible differences in marker profiles of responders and non-responders, as potential marker leads. The top leads representing alternative 3D genomic conformations were then translated into a qPCR format, evaluated and reduced to a molecular classifier. The classifier was then validated on samples from the observational trial and from independent validation cohorts. Here we report on the development of the 3D genomic biomarker panel with clinical utility in predicting response to immune checkpoint inhibitors targeting PD-(L)1 across a variety of oncological indications. These biomarkers reflect prevalent regulatory settings at the level of 3D genomics in the dynamic equilibrium with the patient immune system. They are systemically present at baseline, prior to treatment and have predictive value for response/non-response outcomes to ICI treatments, either with a PD-1 or PD-L1 monoclonal antibody antagonist.

## Materials and Methods

### Patient characteristics

Whole blood samples, 181 in total, were obtained from consented patients enrolled in observational trial “Identifying and Developing Chromosomal Conformation Signatures in Patients Undergoing Cancer Immunotherapy” at Mount Miriam Cancer Hospital (MMCH) in Penang, Malaysia. Additionally, 48 samples were procured commercially (**Supplemental Table 1**). Thirty-two (32) patients were used in *EpiSwitch*® screening and discovery stage, 77 in training model cohort, 24 and 128 patients in two independent validation cohorts. The subject pool represented a multinational set of ICI treated cases, with over 40 distinct oncological diagnoses, from the United States, Europe, and Asia. Patient indications, treatments, clinical outcomes, calls by *EpiSwitch*® classifier, sample use in different stages of the biomarker development are listed in (**Supplemental Table 1)**. Disease response or progression to the therapy was assessed by the investigators according to RECIST 1.1 guidelines [53].

### Preparation of 3D genomic templates

*EpiSwitch*^*®*^ 3D libraries, with chromosome conformation analytes converted to sequence-based tags, were prepared from frozen whole blood samples. using EpiSwitch^®^ protocols following the manufacturer’s instructions for *EpiSwitch*^*®*^ Explorer Array kits (Oxford BioDynamics Plc), samples were processed on the Freedom EVO 200 robotic platform (Tecan Group Ltd). Briefly, 50 mL of whole blood sample was diluted and fixed with a formaldehyde containing EpiSwitch buffer. Density cushion centrifugation was used to purify intact nuclei. Following a short detergent-based step to permeabilise the nuclei, restriction enzyme digestion and proximity ligation were used to generate the 3D libraries. Samples were centrifuged to pellet the intact nuclei before purification with an adapted protocol from the QIAmp DNA FFPE Tissue kit (Qiagen) Eluting in 1x TE buffer pH7.5. 3D libraries were quantified using the Quant-iT™ Picogreen dsDNA Assay kit (Invitrogen) and normalised to 5 ng/ml prior to interrogation by PCR.

### Array design

Custom microarrays were designed using the *EpiSwitch*^*®*^ pattern recognition algorithm, which operates on Bayesian-modelling and provides a probabilistic score that a region is involved in long-range chromatin interactions. The algorithm was used to annotate the GRCh38 human genome assembly across ∼1.1 million sites with the potential to form long-range chromosome conformations [29–36]. The most probable interactions were identified and filtered on probabilistic score and proximity to protein, long non-coding RNA, or microRNA coding sequences. Predicted interactions were limited to *EpiSwitch*^*®*^ sites greater than 10 kb and less than 300 kb apart. Repeat masking and sequence analysis was used to ensure unique marker sequences for each interaction. The *EpiSwitch*^*®*^ *Explorer* array (Agilent Technologies, Product Code X-HS-AC-02), containing 60-mer oligonucleotide probes was designed to interrogate potential 3D genomic interactions. In total, 964,631 experimental probes and 2,500 control probes were added to a 1 × 1 M CGH microarray slide design. The experimental probes were placed on the design in singlicate with the controls in groups of 250. The control probes consisted of six different *EpiSwitch*^*®*^ interactions that are generated during the extraction processes and used for monitoring library quality. A further four external inline control probe designs were added to detect non-human (*Arabidopsis thaliana*) spike in DNA added during the sample labelling protocol to provide a standard curve and control for labelling. The external spike DNA consists of 400 bp ssDNA fragments from genomic regions of *A. thaliana*. Array-based comparisons were performed described previously, with the modification of only one sample being hybridised to each array slide in the Cy3 channel [46]

### Translation of array-based 3D genomic markers to PCR readouts

The top array-derived markers identified in our previous study were interrogated using OBD’s proprietary primer design software package to identify genomic positions suitable for a hydrolysis probe based real time PCR (RT-PCR) assay [47]. Briefly, the top array-derived markers associated with predictive potential to differentiate between response and non-response to ICI outcomes were filtered on logistic regression Glmnet coefficient. PCR primer probes were ordered from Eurofins genomics as salt-free primers. The probes were designed with a 5’ FAM fluorophore, 3’ IABkFQ quencher and an additional internal ZEN quencher and ordered from iDT (Integrated DNA Technologies) [55]. Each assay was optimised using a temperature gradient PCR with an annealing temperature range from 58-68° C. Individual PCR assays were tested across the temperature gradient alongside negative controls including soluble and unstructured commercial TaqMan human genomic DNA control (Life Technologies) and used a TE buffer only negative control. Assay performance was assessed based on Cq values and reliability of detection and efficiency based on the slope of the individual amplification curves. Assays that passed the quality criteria and presented with reliable detection differences between the pooled samples associated with responders and non-responders to ICI treatment outcomes were used to screen individual patient samples.

### *EpiSwitch®* PCR

Each patient sample was interrogated using RT-PCR in triplicate. Each reaction consisted of 50 ng of *EpiSwitch®* library template, 250 mM of each of the primers, 200 mM of the hydrolysis probe and a final 1X Kapa Probe Force Universal (Roche) concentration in a final 25 ml volume. The PCR cycling and data collection was performed using a CFX96 Touch Real-Time PCR detection system (Bio-Rad). The annealing temperature of each assay was changed to the optimum temperature identified in the temperature gradients performed during translation for each assay. Otherwise, the same cycling conditions were used: 98°C for 3 minutes followed by 45 cycles of 95°C for 10 seconds and 20 seconds at the identified optimum annealing temperature. The individual well Cq values were exported from the CFX manager software after baseline and threshold value checks. All Cq values obtained for individual samples and markers are available online (*https://github.com/oxfordBiodynamics/medrxiv/tree/main/CiRT%20publication*) A total of 20 3D genomic markers that passed the translation phase were screened on 32 responder and non-responder samples as a marker reduction step based on statistical criteria to identify the top 8 discriminating markers. These markers were evaluated on 78 individual samples from the training cohort as part of classifier model design. They were then used to screen the independent validation cohorts of 24 and 128 samples.

### Genomic mapping

The 24 3D genomic markers from the statistically filtered list with the greatest and lowest abundance scores were selected for genome mapping. Mapping was carried out using Bedtools closest function for the 3 closest protein coding loci – upstream, downstream and within the long-range chromosome interaction (Gencode v33). All markers were visualized using the *EpiSwitch®* Analytical Portal.

### Statistical analysis

The 20 markers screened on 32 individual patient samples in Screen 1 and 2 were subject to permutated logistic modelling with bootstrapping for 500 data splits and non-parametric Rank Product analysis (EpiSwitch® RankProd R library). Two machine learning procedures (eXtreme Gradient Boosting: XGBoost and CatBoost) were used to further reduce the feature pool and identify the most predictive/prognostic, 3D genomic markers. The resulting markers were then used to build the final classifying models using CatBoost on 78 sample cohort. Catboost is a member of the Gradient Boosted Decision Trees (GBDT’s) machine learning ensemble techniques [56]. All analysis was performed using R statistical language with Caret, XGBoost, SHAPforxgboost and CatBoost libraries.

### Biological network/pathway analysis

Protein interaction networks and pathway enrichment were generated using the Search Tool for the Retrieval of Interacting proteins (STRING) and Reactome Pathway Browser databases [57–59].

## Results

### Whole genome array profiling for discovery of predictive 3D genomic marker leads in baseline Immuno-Oncology (IO) patients at baseline

The *EpiSwitch®* array platform was used for whole genome screening and discovery of 3D genomic biomarker leads. It has been utilized to date on over 120 IO patients, generating over 104 million individual chromosome conformation data points. We based our initial selection of marker leads on the screening results from whole genome EpiSwitch array for 32 patients from the observational trial at Mount Miriam Cancer Hospital (MMCH). These patients were treated with either Pembrolizumab, Atezolizumab or Durvalumab, and were diagnosed with one of the following indications: lung cancer, kidney cancer, nasopharyngeal cancer, sagittal sinus carcinoma, neuroendocrine tumour, vulvar carcinoma. Among the responders, those patients were confirmed in the durable nature of their response and absence of acquired resistance.

From over 30 million data points, following the logistic regression Glmnet coefficient selection for baseline responders and non-responders we identified the top 72 marker leads associated with predictive value for response and non-response to ICI (Table 1).

**Table 1.**
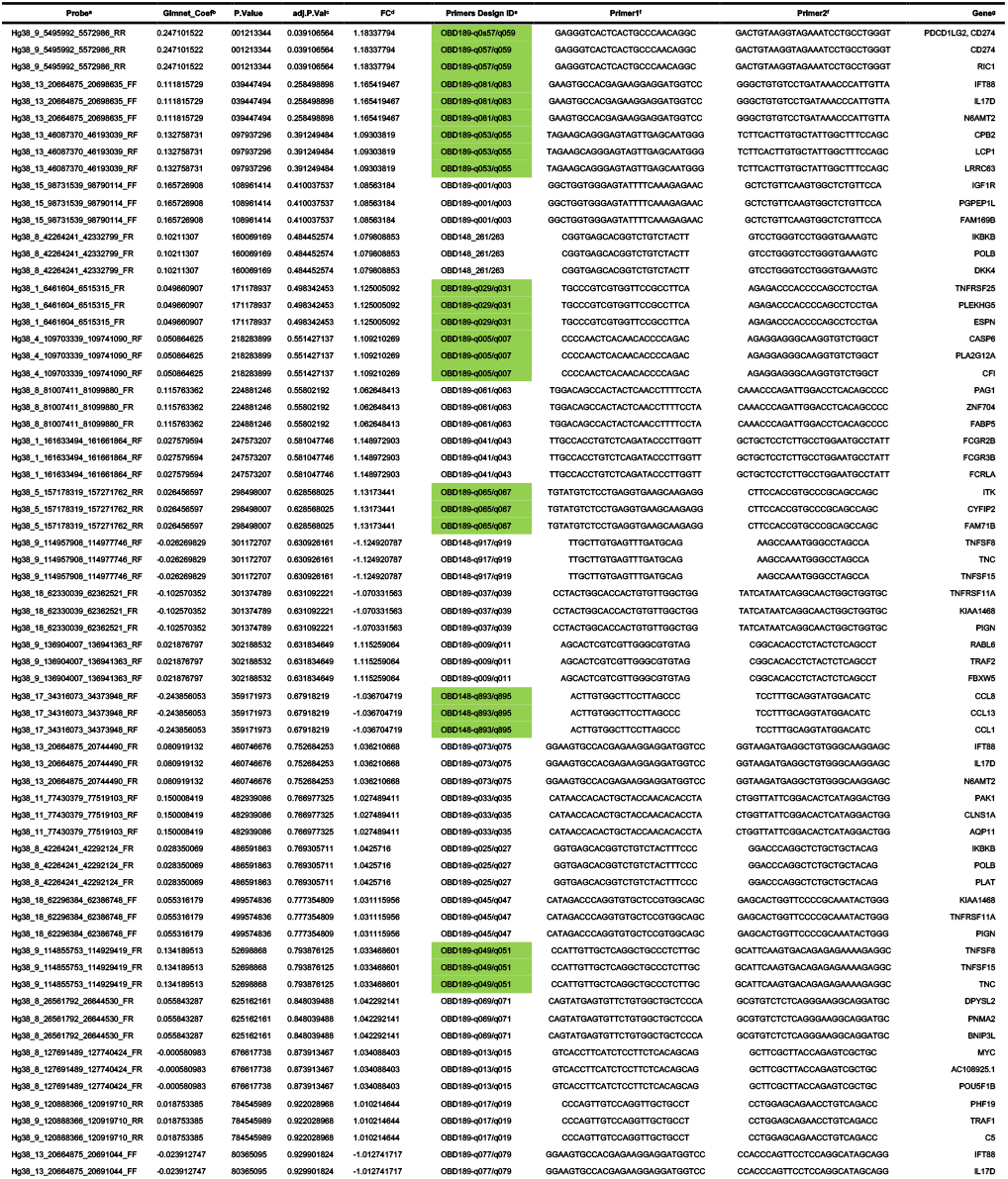
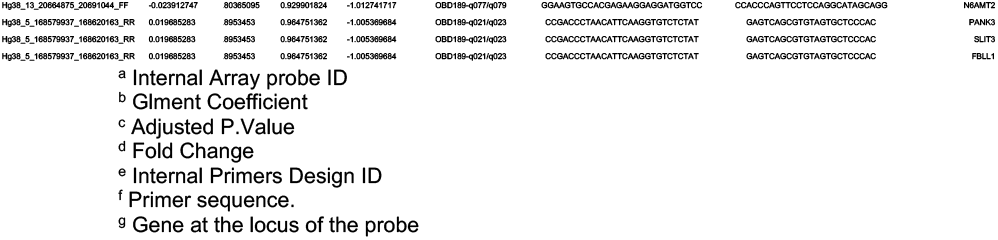
Array based 3D genomic marker leads for baseline response to ICI. Top array-based 3D genomic markers identified from over 30 million data points in 32 responders and non-responders across several indications and several choices of ICI: Pembrolizumab, Atezolizumab or Durvalumab. Probe – array-based marker coordinates for long-range interaction junction; Glmnet Coefficient, P value, Adjusted P value and Fold Change (FC) are array-based measures of markers in comparison of responders and non-responders groups; Primer Design ID – qPCR primer probes designs corresponding to array probes. 24 optimal in silico designs (marked in green) were taking forward to quality control checks in temperature gradient analysis. Gene – identity of genes of interest in the location of 3D genomic markers.

It is important to point out that the most significant marker in this selection was associated with CD274 and PDCD1LG2 loci, at the junction of the genes encoding for PD-L1 and PD-L2 checkpoint inhibitors. Functionally this suggests a regulatory event associated with the predictive profile for response to ICI and leading to specific conditional differences among patients as captured systemically through regulatory 3D genomic profiles. This is consistent with earlier observations that PD-L1 expression levels, as reflected in HIT testing, could share predictive values under specific conditions [27,28].

### Identification of the top predictive 3D genomic markers for ICI treatment outcomes

Following the establish methodology for *EpiSwitch®* marker reduction [43,47], we employed a stepwise approach to translate array-based marker leads and identify a minimal set of biomarkers for predictive stratification of ICI treatment response outcome (Figure 1). From over 30 million data points on array profiling, the top 72 markers were used for translation into qPCR format. The design and sequencing restrictions on the primers and fluorescent probes corresponding to the array probe sites of genome long-ranged junction points, has reduced 72 markers to 24 qPCR designs (denoted in Table 1). At the experimental stage of temperature gradient optimization 20 qPCR marker designs have passed quality control (Table 2).

**Table 2.**
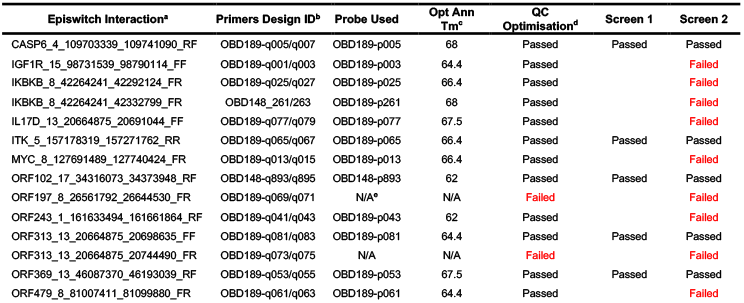
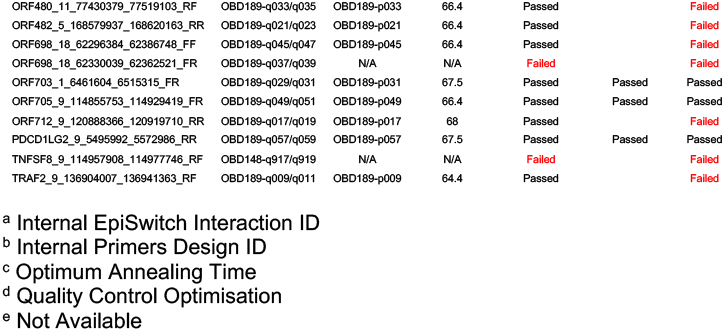
Translation and selection of top markers into qPCR format. Temperature gradient optimization for primer probe designs of the top 24 markers. Markers that failed quality control and feature reduction in Screen 1 and Screen 2 are marked. EpiSwitch interaction – marker position; Primer Design ID and Probe Used – prier and probe identities; Opt Ann Tm – optimized annealing T for detection; QC Optimization – temperature gradients testing; Screen 1 – selection on pooled groups of responders and non-responders; Screen 2 – final selection on the basis of individual patients profiling.

**Figure 1.**
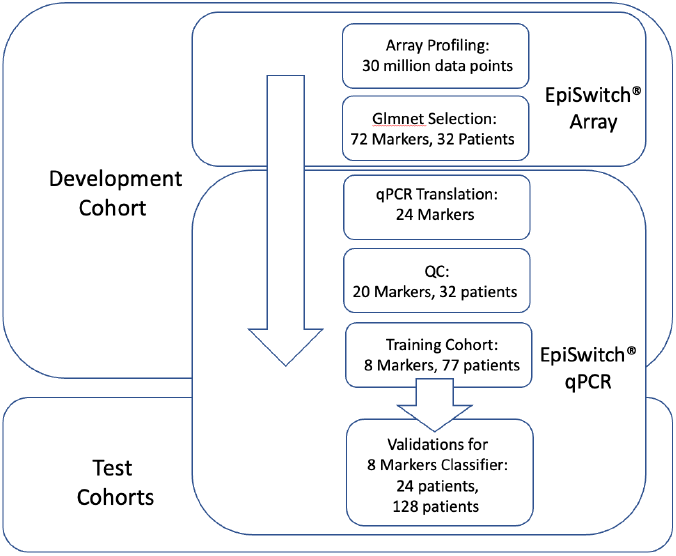
Workflow for development and testing the 3D genomic classifier model for prediction of ICI treatment response outcome. From EpiSwitch array screening profiles accounting for over 30 million data points, 72 top array markers were selected based on Glmnet logistic regression. 24 markers qualified for translation into EpiSwitch qPCR format, of which 20 markers have passed quality control and feature reduction control on 32 patient samples. Training cohort of 77 patients was used to build predictive classifier model based on 8 qPCR marker. It was then validated on independent cohorts of 24 and 128 patients.

All 20 markers then underwent feature reduction in several steps (Materials and Methods). Firstly, they were evaluated in Screen 1 on pooled samples from 32 patients representing either responders, non-responders or stable disease. These sample cohorts represented patients treated with Pembrolizumab, Atezolizumab and Durvalumab (marked in Supplemental Table 1). Based on the results from screen 1 the top 13 markers were advanced to screen 2, where 13 markers were run on 26 patients individually, further reducing the selection to top 8 markers (Table 2).

The 8 selected markers were further evaluated on the training cohort of 77 patients (see Supplemental Table 1). Blood samples taken from patients prior to initiation of ICI therapy were used together with clinical assessment for response status according to RECIST 1.1 criteria. Baseline clinical characteristics were similar between responders and non-responders (Supplemental Table 1). The training cohort of 77 patients represented ICI treatments: Pembrolizumab, Atezolizumab and Durvalumab, with cancers from lung, pancreas, bladder, kidney, larynx, salivary gland, liver, breast, colon, nasopharynx, meninges, and vulva.

We used these 8 markers to generate the 3D genomic classifier with predictive ability for ICI response, which we then applied to the independent test cohorts.

### Testing of the predictive 3D genomic biomarker panel for response to ICI treatments on independent patient cohorts

To access the predictive power of the classifier model, the 8-marker 3D genomic panel was validated on an independent baseline Test cohort #1 (Supplemental Table 1). No samples from that cohort were used to build and refine the model. Samples were collected at the Mount Miriam Cancer Hospital observational study and shipped to OBD’s processing facility in Oxford, UK. The EpiSwitch platform readouts for the eight-marker classifier model were uploaded to the EpiSwitch Analytical Portal for analysis. Clinical outcomes for the test cohort #1 included a balanced representation of 12 responders and 8 non-responders and 3 stable diseases. It is important to mention that from the start of the model classification design all the stable disease cases were considered to be non-responders. EpiSwitch predictive calls based on 8-marker model demonstrated high performance of 83% balanced accuracy and 83% positive predictive value in the test cohort #1 (Figure 2A). Across all 101 patients used in this study in both training and testing cohorts, the test demonstrated positive predictive value 96% and balanced accuracy of 96% (Figure 2B). The patients represented ICI treatments: Pembrolizumab, Atezolizumab and Durvalumab, and indications including cancer from cervix, kidney, liver, lung, neuroendocrine, meninges vulva.

**Figure 2.**
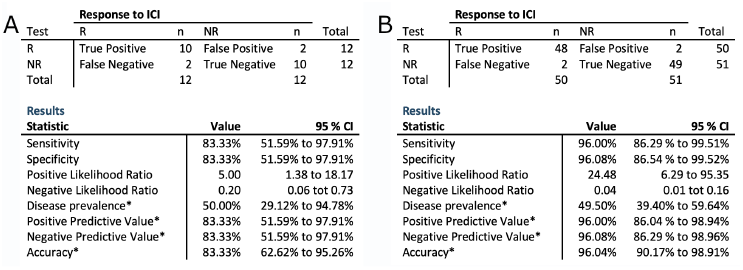
Performance of the prognostic biomarker classifier for calling response (R) and non-response (NR) to ICI treatment outcome. (A)Confusion matrix and test performance statistics for the 8-marker classifier on the 24 patients of the test cohort and (B) in the combined Train and Test cohorts of 102 patients. Note: (*) These values are dependent on disease prevalence.

We have further extended our validation exercise, obtaining additional 128 samples as Test cohort #2 (Supplemental Table 1). In contrast to the balanced testing cohort #1, these 128 base line patients represented largely non-responder group, with only 9 responders present. EpiSwitch predictive calls based on 8-marker model demonstrated 76% accuracy, 78% sensitivity and 76% specificity on 128 patients in the test cohort #2 (Figure 3A). Across all 229 patients used in this study, with 170 non-responders and 59 responders in training and both testing cohorts, the test demonstrated accuracy of 85%, sensitivity 93%, specificity 82%, with positive predictive value 64%, and negative predictive value 97% (Figure 3B). The combined cohort of patients represented treatments with anti-PD-1 or anti-PD-L1 ICI therapies, including: Pembrolizumab, Atezolizumab, Durvalumab, in a variety of indications including cancer from: pancreas, soft tissue (alveolar soft part sarcoma), bile duct (cholangiocarcinoma), bladder, cervix, vulva, kidney, larynx, parotid gland (mucoepidermoid carcinoma), colon, liver, breast, lung (adenocarcinoma, small cell carcinoma, squamous cell carcinoma), lymphoepithelial carcinoma, prostate, stomach, high-grade neuroendocrine tumor, melanoma, nasopharynx, meninges, oral cavity, brain, and maxilla.

**Figure 3.**
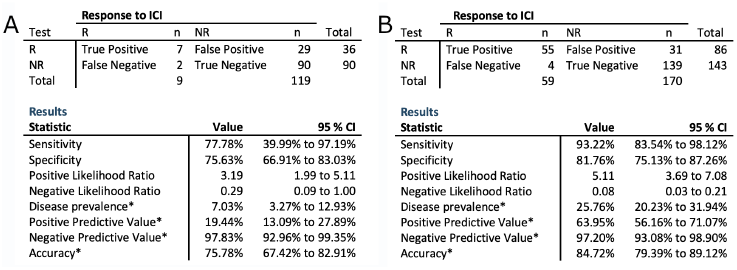
Performance of the prognostic biomarker classifier for calling response (R) and non-response (NR) to ICI treatment outcome. (A) Confusion matrix and test performance statistics for the 8-marker classifier on the 128 patients of the test cohort and (B) in the combined Train and Test cohorts of 240 patients. Note: (*) These values are dependent on disease prevalence

## Discussion

Cancer treatment has been revolutionized by the development of therapies that target the immune checkpoint response [3]. However, only a minority of patients benefit from ICI therapies today. With high prevalence of immune-related adverse events accompanying ICI treatments, clinical decisions and evaluation of risk-benefit ratio would greatly benefit from the development of molecular biomarkers to predict clinical response.

Here we used a 3D genomics biomarker approach, which has already demonstrated successful development of valuable prognostic and predictive biomarkers in oncology and autoimmune conditions [52,60,61]. We have identified 8 systemic 3D genomic biomarkers, that when assessed as a molecular classifier in blood samples from a diverse group of baseline patients treated with anti-PD-(L)1 immune checkpoint inhibitors, gave an early readout of likely response or non-response to ICI therapies prior to treatment initiation. Importantly, as a reflection of the network regulation, the identified 3D genomic markers are associated with genes related to the regulation of the immune system (Figure 4). This is consistent with the concept that ICI therapies exert their therapeutic effects on tumour cells by enhancing the cell-mediated immune response [3]. The captured 3D genomic differences observed between responder and non-responder baseline patients also identified 3D biomarkers corresponding to genetic loci and pathways (Figure 5) including NF-kB and TGF-b, whose biologic functions are related to checkpoint response [62–64]. The top 3D genomic markers identified by the EpiSwitch Explorer Array profile as associated at baseline with response/non-response to ICI treatment were also analysed using the Search Tool for Retrieval of Interacting Genes (STRING) database. The view from the established protein-protein networks confirmed that the coding regions associated with EpiSwitch top 24 marker leads are highly connected (Figure 6). In fact, no additional nodes were added to generate the network based on the 24 EpiSwitch marker leads. This is consistent with the regulatory network formed by the 3D genome architecture, as an integrator of molecular multi-omics mechanisms [30] and being concordant with controls of the protein expression and cellular phenotype.

**Figure 4.**
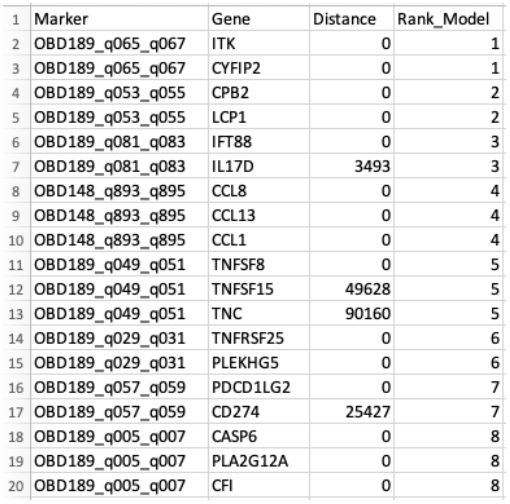
Mapping of the top 8 predictive 3D genetic markers. Genes – genes associated with location of the 8 top 3D genomic markers analyzed in this study. Distance – actual distance of the 3D long-range interaction marker from the gene ORF. Rank_Model – ranking order of the markers in the classifier model.

**Figure 5.**
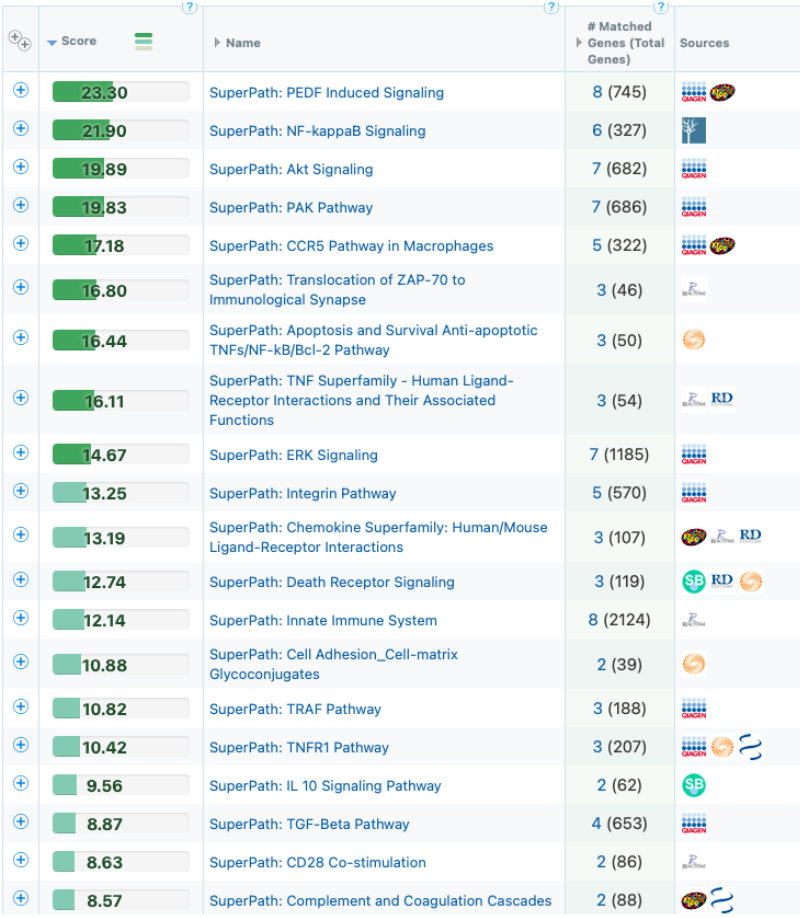
Mapping of the top 8 predictive 3D genomic markers to biological pathways. Analysis of the top 8 3D genomic markers separating PD-(L)1 ICI responders and non-responders at baseline.

**Figure 6.**
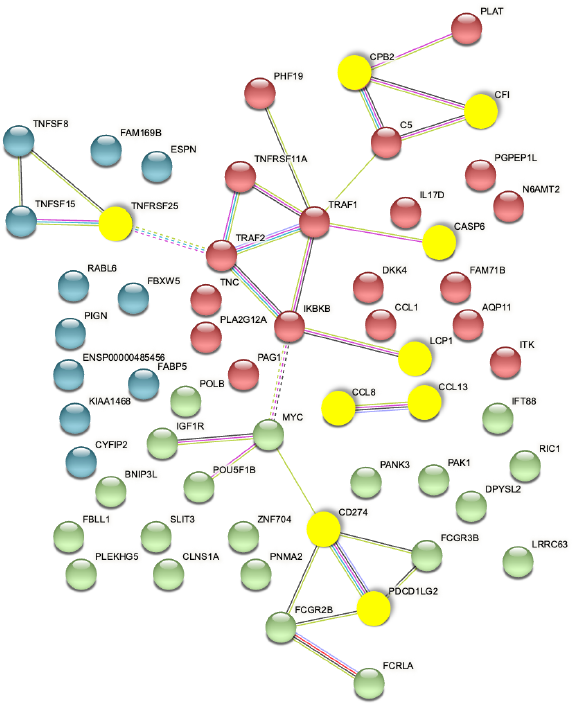
STRING Network associated with baseline prediction of response /non-response to ICI treatment. The proteins encoded by genes in the top 3D genomic markers associated with baseline response/non-response to ICI profile show a highly connected network. The yellow nodes mark the coding regions associated with the top 8 markers of the validated classifier.

Among other interesting observations is the high prevalence of the validated 3D marker responsible for conditional regulatory long-range interactions in the region spanning CD274(PD-L1) and PDCD1LG2 (PD-L2) (Figure 7). In fact, this systemic predictive marker has been consistently observed across multiple cohorts of IO patients, indicating that the complex network regulation manifestly shares predictive values both at the systemic 3D genomic level and at the level of PD-L1 gene expression in established IHC testing [3]. The EpiSwitch Portal view of this marker (Figure 7) identifies variable allelic frequency SNPs and the affected transcriptional factor (TF) binding motifs around the marker anchor sites. Three TFs were identified at the proximal anchor site of the marker: NFIC, ZEB1, MZF1. Analysis of GeneHancer repository (GeneCards) shows evidence of enhancer controls (Gene association score) over CD274 (PD-L1) for the distal anchor site of the same marker. All together this strongly suggests that the regulatory effect of the EpiSwitch marker over the PD-L1 locus is executed through coordinated activities of the CD274 enhancer and ZEB1 TF activities, well in line with other independent studies [65,66].

**Figure 7.**
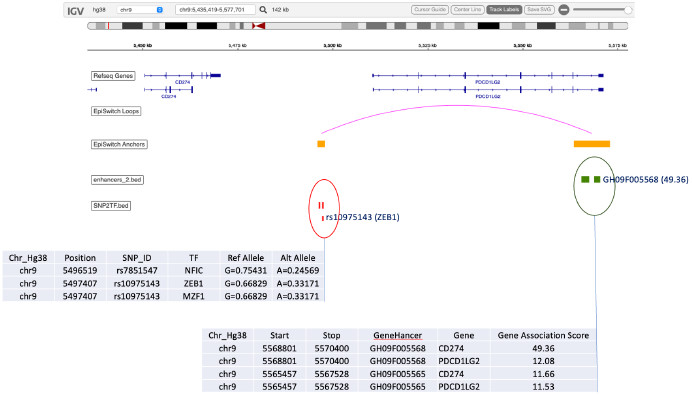
EpiSwitch Data Portal view of the EpiSwitch Marker located in CD274/PDCD1LG2 region. Anchor sites brought into juxtaposition by chromosome long range interactions of the EpiSwitch marker, overlap with the listed SNPs and enhancers. Analysis of SNPs identified affected binding sites for transcription factors (TFs) such as ZEB1 [65]. GeneCards analysis of the enhancers identifies CD274 enhancer at the distal anchor site of the EpiSwitch marker. CD274 corresponds to PD-L1, and PDCD1LG2 to PD-L2 immune checkpoints.

Altogether, this indicates that the predictive model developed in this study has high biological relevance and is anchored on baseline presence of a prevalent systemic 3D genomic profile, functionally linked to immuno-genetic settings conducive to clinical outcome of response to PD-(L)1 blockade. The systemic nature of the observed marker profile is not surprising since the 3D genomic approach described here was done on whole blood samples, with dominant lymphocytic representation. The fact that some T-cell related loci were found to bear conditional regulatory differences in 3D genome architecture, in association with clinical phenotypes of responder/non-responder profiles, is a fascinating regulatory phenomenon. It is consistent with multiple observations of the role 3D genome architecture plays in regulation of oncological phenotype and clinical outcomes [30].

A simple blood-based assay that provides a readout of likely response to PD-(L)1 ICI therapy could be a valuable asset for oncologists considering ICI therapy, since only a minority of patients experience durable tumour responses. For example, ORR for metastatic NSCLC is 24% and 16%, for treatment-naïve and previously treated patients, respectively [67]. A positive PD-L1 expression result from an immunohistochemistry (IHC) test, which increases ORR to 39.7% (PD-L1 50% or greater) [67]. Many of the remaining patients derive no clinical benefit, suffering significant drug-related adverse events (∼14%) or even death from pulmonary toxicity (>1%) [68]. Moreover, approximately 15% of non-responders may also be at risk of hyper-progressive disease (HPD), accelerated by ICI therapy and shortening their lives by months [69]. Today, despite a low response rate, many patients are prescribed ICI therapies at a substantial financial cost to both payers and patients [70]. All together, these aspects highlight the importance of a response predictive assay to improve patient selection for optimized treatment, better overall treatment-decision planning, potential utilization of alternative effective treatments, and avoiding futile care and unnecessarily toxicity, and efficiently managing costs and resources [70].

The current study represents a proof-of-concept that 3D genomic changes, measurable in blood, can be used as biomarkers of response prediction to PD-(L)1 ICIs in oncology. In this study all patients underwent ICI treatment, with no comparator arm undergoing control treatment or basic standard of care. The single-arm design of this study does not allow us to definitively differentiate between predictive and prognostic values of the classifier. Extension of this work to a larger number of advanced patients with different ICI therapies and with a comparator treatment arm, could help further validate the predictive value of the developed EpiSwitch ICI biomarker classifier. A pan-therapy application of the test with response prediction across tumour types with historically low ORRs could help to improve both patient outcomes and increase the cost effectiveness of cancer care [71,72].

## Conclusions

With the rapid advancement of novel therapies targeting the immune checkpoint pathway, there is a pressing need to develop better biomarkers to assess likely clinical response in advance of therapeutic intervention. Here we report on a novel 3D genomics approach to identify predictive blood-based markers that can identify, with high accuracy, individuals that are likely to respond to PD-(L)1 ICIs monotherapy across tumour types with low ORRs. The 3D genomics approach described here has been developed into a clinical assay to assist in treatment decisions, help improve patient selection for optimized treatment, help better utilize alternative effective treatments, minimize or avoid unnecessarily toxicity, and efficiently manage costs and resources.

## Data Availability

The data that support the findings of this study are included in this published article, it's supplementary files and on the following Github repo:

https://github.com/oxfordBiodynamics/medrxiv/tree/main/CiRT%20publication

## Declarations

*EH, MD, CK, AW, FCS, MS, RP, AD, BE, MP, AR, and AA are full-time employees at Oxford BioDynamics plc and have no other competing financial interests*.

## Author Contributions

*EH, AA conceived the study, SN, CRL, CSK, ATS, RR, HKF, FLWL assisted with study design and clinical samples collection. AD, MS, EH and AR assisted with EpiSwitch*^*®*^ *array design. JWM, AB and ZL assisted with design of experiments. MD and AA planned and reviewed experiments. CK, AW, FCS and EH analysed the data. MD, RP, AD, MP, BE performed experiments, with support from MT. JWW, RH, JL helped with interpretation of the data and writing of the manuscript. JWW, MD, EH, and AA. wrote and reviewed the manuscript*.

## Acknowledgements

*The authors would like to thank Professor Jane Mellor, University of Oxford, for helpful discussions; members of OBD Louis James, Thomas Lavin, Catriona Williams, Aemilia Katzinski, Paulina Braier, Pieter Harvey, and Kalsoom Rana for their operational support, Olly Hunter for assistance in data analysis. In addition, we acknowledge Agilent Technologies, Inc. for supply of EpiSwitch*^*®*^ *designed CGH microarray slides*.

## Consent for publication

Written informed consent for publication was obtained from all authors.

## Availability of data and materials

The datasets used and/or analysed during the current study are available from the corresponding author on reasonable request.

## Ethics approval and consent to participate

All patients signed informed consent forms prior to providing blood samples. All ethical guidelines were followed.

